## Supplementary Table 1 for "Long Covid stigma: estimating burden and validating scale in a UK-based sample"

Table 1 Sample characteristics (n=1166)

|  | n | % |
| --- | --- | --- |
| **Age (mean 47.7, SD 10.6)** |  |  |
| 18-30 | 63 | 5.5 |
| 31-45 | 415 | 36.0 |
| 46-59 | 519 | 45.1 |
| ≥60 | 155 | 13.5 |
| Missing | 14 | 1.2 |
| **Gender** |  |  |
| Male | 173 | 15.0 |
| Female | 965 | 83.8 |
| Non-binary or other | 14 | 1.2 |
| Missing | 14 | 1.2 |
| **Ethnicity** |  |  |
| White | 1096 | 95.4 |
| Minority ethnic groups | 53 | 4.6 |
| Missing | 17 | 1.5 |
| **Country of residence** |  |  |
| UK – England | 778 | 67.3 |
| UK – Scotland | 111 | 9.6 |
| UK – Wales | 58 | 5.0 |
| UK – Northern Ireland | 9 | 0.8 |
| Outside the UK | 200 | 17.3 |
| Missing | 10 | 0.9 |
| **Educational** **qualification** |  |  |
| No formal qualifications | 13 | 1.1 |
| O levels or equivalent | 97 | 8.3 |
| A levels or equivalent | 151 | 13.0 |
| University degree or above | 902 | 77.5 |
| Other | 1 | 0.1 |
| Missing | 2 | 0.2 |
| **Duration of illness** |  |  |
| <12 months | 40 | 3.5 |
| 12-<15 months | 66 | 5.8 |
| 15-<18 months | 51 | 4.5 |
| >18 months | 989 | 86.3 |
| Missing | 20 | 1.7 |
| **Employment status** |  |  |
| Employed | 780 | 67.0 |
| Unable to work | 341 | 29 |
| Student/Volunteer | 25 | 2.2 |
| Unemployed and looking for work | 19 | 1.6 |
| Missing | 1 | 0.1 |
| **Clinical diagnosis of Long Covid received or on health record** |  |  |
| No | 53 | 4.8 |
| Not sure | 129 | 11.7 |
| Have test confirmation of initial Covid infection but no/not sure clinical diagnosis of Long Covid | 73 | 6.6 |
| No official diagnosis but doctors suspect I have Long Covid | 308 | 27.9 |
| Yes, Long Covid as a diagnosis on health record | 540 | 49.0 |
| Missing | 63 | 5.4 |
| **Long Covid Stigma Scale (LCSS) score**, mean (SD) | 20.5 ± 10.7 | |
| Missing | 99 | 8.5 |
| **Disclosure concerns**, mean (SD) | 2.9 ± 2.3 | |
| Missing | 71 | 6.1 |
| **PHQ-8 score**, mean (SD) | 9.2 ± 5.8 | |
| Missing | 103 | 8.8 |

Table 2 Response option frequencies for each stigma item

|  | Full sample  (n=1067) | | | | | Clinical diagnosis  (n=516) | | | | | No clinical diagnosis/unsure  (n=543) | | | | |
| --- | --- | --- | --- | --- | --- | --- | --- | --- | --- | --- | --- | --- | --- | --- | --- |
| Response options* | 0 | 1 | 2 | 3 | 4 | 0 | 1 | 2 | 3 | 4 | 0 | 1 | 2 | 3 | 4 |
| **Enacted stigma items** |  | | | | |  | | | | |  | | | | |
| Because of my illness, some people seemed uncomfortable with me | 27.5 | 20.4 | 37.6 | 13.3 | 1.3 | 18.8 | 20.5 | 44.3 | 14.5 | 1.9 | 35.4 | 20.2 | 31.8 | 11.9 | 0.7 |
| Because of my illness, some people were unkind to me | 48.2 | 21.8 | 22.8 | 7.0 | 0.3 | 40.7 | 24.0 | 27.4 | 7.7 | 0.2 | 55.2 | 19.6 | 18.7 | 6.2 | 0.4 |
| People I care about stopped contacting me after learning I have Long Covid | 54.5 | 17.1 | 19.3 | 8.4 | 0.8 | 43.4 | 19.4 | 25.4 | 11.2 | 0.6 | 64.9 | 15.0 | 13.4 | 5.6 | 1.1 |
| People have acted as if I am dishonest since I have had Long Covid | 45.6 | 20.2 | 23.3 | 9.3 | 1.6 | 42.1 | 22.5 | 23.6 | 10.9 | 0.9 | 48.9 | 17.9 | 23.3 | 7.8 | 2.2 |
| I have been treated with less respect than other people are because of Long Covid | 48.1 | 20.0 | 21.5 | 9.6 | 0.8 | 40.0 | 22.3 | 24.4 | 12.2 | 1.1 | 55.3 | 18.1 | 18.8 | 7.2 | 0.5 |
| **Internalised stigma items** |  |  |  |  |  |  |  |  |  |  |  |  |  |  |  |
| I have felt embarrassed about my illness | 27.8 | 12.1 | 30.5 | 21.4 | 8.2 | 21.9 | 12.2 | 32.0 | 24.1 | 9.9 | 32.7 | 12.3 | 29.6 | 19.0 | 6.5 |
| I have felt embarrassed because of my physical limitations | 16.1 | 9.4 | 29.5 | 31.3 | 13.8 | 11.1 | 8.1 | 28.0 | 36.3 | 16.5 | 20.0 | 10.8 | 31.2 | 27.0 | 11.0 |
| I feel that I have been tainted by Long Covid and am of less value than others because of it | 27.1 | 16.3 | 26.7 | 19.1 | 10.8 | 18.4 | 15.0 | 28.0 | 23.8 | 14.8 | 34.8 | 17.8 | 25.8 | 15.0 | 6.7 |
| I have felt like I am very different from other people on account of Long Covid | 17.5 | 14.9 | 32.2 | 21.9 | 13.5 | 11.3 | 12.8 | 31.4 | 27.6 | 16.9 | 23.1 | 16.8 | 33.0 | 16.8 | 10.3 |
| **Anticipated stigma items** |  |  |  |  |  |  |  |  |  |  |  |  |  |  |  |
| Many people tend to think Long Covid isn’t a real illness | 7.6 | 12.8 | 36.2 | 33.5 | 10.0 | 6.9 | 13.6 | 37.6 | 31.0 | 10.8 | 7.9 | 12.1 | 34.7 | 35.9 | 9.4 |
| I feel that some people assume that having Long Covid is a sign of personal weakness | 18.1 | 15.9 | 34.6 | 23.9 | 7.6 | 12.8 | 15.0 | 36.7 | 26.3 | 9.2 | 23.0 | 16.6 | 32.4 | 21.9 | 6.2 |
| I worry that people with Long Covid lose their jobs when their employers find out | 19.3 | 14.6 | 36.5 | 22.1 | 7.5 | 13.2 | 13.5 | 37.4 | 26.9 | 9.0 | 24.6 | 15.7 | 36.2 | 17.5 | 6.0 |
| I worry that people may judge me negatively when they learn I have Long Covid | 21.4 | 17.6 | 32.8 | 19.5 | 8.7 | 13.8 | 16.8 | 34.8 | 23.0 | 11.6 | 28.4 | 18.4 | 31.1 | 16.1 | 6.0 |
| *Response options indicate 0:Never; 1:Rarely; 2:Sometimes; 3:Often; 4:Always | | | | | | | | | | | | | | | |

Table 3 Factor loadings of individual stigma items on subscales of internalised, enacted and anticipated stigma using confirmatory factor analysis

|  | Full sample  (n=1067) | | | Clinical diagnosis  (n=516) | | | No clinical diagnosis/unsure  (n=543) | | |
| --- | --- | --- | --- | --- | --- | --- | --- | --- | --- |
|  | Enacted | Internalised | Anticipated | Enacted | Internalised | Anticipated | Enacted | Internalised | Anticipated |
| Because of my illness, some people seemed uncomfortable with me | 0.78 |  |  | 0.75 |  |  | 0.79 |  |  |
| Because of my illness, some people were unkind to me | 0.79 |  |  | 0.80 |  |  | 0.78 |  |  |
| People I care about stopped contacting me after learning I have Long Covid | 0.70 |  |  | 0.66 |  |  | 0.71 |  |  |
| People have acted as if I am dishonest since I have had Long Covid | 0.76 |  |  | 0.77 |  |  | 0.77 |  |  |
| I have been treated with less respect than other people are because of Long Covid | 0.86 |  |  | 0.86 |  |  | 0.86 |  |  |
| I have felt embarrassed about my illness |  | 0.76 |  |  | 0.74 |  |  | 0.76 |  |
| I have felt embarrassed because of my physical limitations |  | 0.75 |  |  | 0.72 |  |  | 0.77 |  |
| I feel that I have been tainted by Long Covid and am of less value than others because of it |  | 0.83 |  |  | 0.82 |  |  | 0.83 |  |
| I have felt like I am very different from other people on account of Long Covid |  | 0.73 |  |  | 0.67 |  |  | 0.76 |  |
| Many people tend to think Long Covid isn’t a real illness |  |  | 0.64 |  |  | 0.70 |  |  | 0.63 |
| I feel that some people assume that having Long Covid is a sign of personal weakness |  |  | 0.77 |  |  | 0.79 |  |  | 0.74 |
| I worry that people with Long Covid lose their jobs when their employers find out |  |  | 0.58 |  |  | 0.51 |  |  | 0.60 |
| I worry that people may judge me negatively when they learn I have Long Covid |  |  | 0.86 |  |  | 0.84 |  |  | 0.87 |
| CFI |  | 0.970 |  |  | 0.975 |  |  | 0.968 |  |
| TLI |  | 0.956 |  |  | 0.963 |  |  | 0.952 |  |
| RMSEA |  | 0.065 |  |  | 0.058 |  |  | 0.068 |  |
| SRMR |  | 0.037 |  |  | 0.040 |  |  | 0.039 |  |
| χ2/df |  | 5.5 |  |  | 2.7 |  |  | 3.5 |  |
| Cronbach’s alpha | 0.88 | 0.86 | 0.82 | 0.87 | 0.84 | 0.81 | 0.89 | 0.86 | 0.82 |

Table 4 Correlations between stigma scores, eight-item Patient Health Questionnaire (PHQ-8 score) and disclosure concerns

|  | Full sample | | | | Clinical diagnosis | | | | No clinical diagnosis/unsure | | | |
| --- | --- | --- | --- | --- | --- | --- | --- | --- | --- | --- | --- | --- |
|  | PHQ-8 score | p-value | Disclosure concerns | p-value | PHQ-8 score | p-value | Disclosure concerns | p-value | PHQ-8 score | p-value | Disclosure concerns | p-value |
| Overall LCSS | 0.46 | <0.001 | 0.62 | <0.001 | 0.45 | <0.001 | 0.65 | <0.001 | 0.46 | <0.001 | 0.62 | <0.001 |
| Enacted stigma subscale | 0.34 | <0.001 | 0.50 | <0.001 | 0.31 | <0.001 | 0.53 | <0.001 | 0.35 | <0.001 | 0.48 | <0.001 |
| Internalised stigma subscale | 0.48 | <0.001 | 0.53 | <0.001 | 0.47 | <0.001 | 0.55 | <0.001 | 0.47 | <0.001 | 0.53 | <0.001 |
| Anticipated stigma subscale | 0.38 | <0.001 | 0.59 | <0.001 | 0.38 | <0.001 | 0.59 | <0.001 | 0.38 | <0.001 | 0.61 | <0.001 |

Table 5 Prevalence of reported stigma

|  | Experienced stigma sometimes or more often | | | | Experienced stigma often/always | | | |
| --- | --- | --- | --- | --- | --- | --- | --- | --- |
|  | Full sample | Clinical diagnosis | No clinical diagnosis/unsure | p-value | Full sample | Clinical diagnosis | No clinical diagnosis/unsure | p-value |
| Overall LCSS | 95.1 | 97.5 | 93.0 | 0.001 | 76.2 | 83.0 | 70.2 | <0.001 |
| Enacted stigma | 63.4 | 71.3 | 56.6 | <0.001 | 25.6 | 29.1 | 22.3 | 0.01 |
| Internalised stigma | 86.8 | 92.2 | 82.1 | <0.001 | 59.3 | 69.1 | 50.4 | <0.001 |
| Anticipated stigma | 90.1 | 93.0 | 88.4 | 0.009 | 59.5 | 63.5 | 56.0 | 0.01 |
